# Can Predictive Modeling Inform the Selection of Time Zero for Target Trial Emulations? An Empirical Study of Atorvastatin Initiation in Medicare Beneficiaries

**DOI:** 10.64898/2026.05.05.26352148

**Authors:** Christopher G. Rowan, Steven M. Brunelli, Camille Maringe

**Affiliations:** Veritas Research Foundation, San Clemente, CA USA; Rutgers University, Center for Pharmacoepidemiology & Treatment Science (PETS), New Brunswick, NJ, USA; Inequalities in Cancer Outcomes Network (ICON), Health Services Research and Policy Department, London School of Hygiene and Tropical Medicine London, UK

**Author notes:** **Corresponding author** Christopher G. Rowan, PhD, 4030 Calle Marlena, San Clemente, CA 92672, https://www.veritasrf.org/.

**Keywords:** Target trial emulation, time zero event, time zero, atorvastatin initiation, Medicare, pharmacoepidemiology, selection bias, elderly

## Abstract

**Purpose:** When emulating trials of medication initiation using real-world data, there may be ambiguity regarding the most suitable time zero event for the research question of interest. The time zero event must be strongly associated with the clinical indication for treatment, confer a reasonably high probability of actual treatment initiation, and be measurable with sufficient temporal precision in the source data. When it is uncertain whether a candidate event will satisfy these three conditions simultaneously, empirical identification of predictors of medication initiation can provide valuable guidance. The objective of this study was to empirically identify predictors of incident atorvastatin initiation to inform the definition of time zero for future target trial emulations.

**Methods:** A retrospective cohort study was conducted using Medicare claims data from January 1, 2018 – December 31, 2019 (study period). The study population included beneficiaries: age ≥ 65 years with ≥ 12 months of continuous enrollment and no statin medication for ≥ 12 months as of the study period start date. The index date was the date of the first pharmacy dispensing claim for a newly initiated medication during the study period, and classified as atorvastatin or non-atorvastatin (non-atorvastatin initiators randomly sampled at 10%). Candidate predictor variables were ascertained within 6 months pre-index and included demographics, comorbidities, healthcare utilization, and pharmacotherapy. A prespecified eight-step procedure was used to identify significant, independent predictors of incident atorvastatin initiation (vs initiating a medication other than atorvastatin).

**Results:** The study cohort comprised 117,069 incident atorvastatin initiators and 393,083 non-atorvastatin initiators. The strongest predictors of atorvastatin initiation were having an inpatient admission for cerebral infarction (OR 11.29, 95% CI 10.47-12.17) and myocardial infarction (OR 6.56, 95% CI 5.92–7.27). For example, a White male with a recent hospitalization for cerebral infarction had a predicted probability of atorvastatin initiation of 69% (95% CI 68-70).

**Conclusion:** The empirically identified predictors of atorvastatin initiation among statin naïve Medicare beneficiaries align with ACC/AHA guidelines recommending prompt statin therapy for secondary prevention. These predictors satisfy the three key requirements for a valid time zero event and should mitigate channeling bias and residual confounding in future target trial emulations.

**KEY POINTS:**

- **Findings**: Acute myocardial infarction and cerebral infarction recorded during an inpatient admission were the strongest predictors of incident atorvastatin initiation among statin-naïve Medicare beneficiaries age 65 years and older.
- **Clinical Context:** These findings align with current American College of Cardiology/American Heart Association guidelines that recommend prompt statin therapy for secondary prevention after these acute cardiovascular events.
- **Implications for Future Research:** Anchoring the time zero event to an inpatient admission for myocardial/cerebral infarction satisfies the three key requirements for a valid time zero event when studying medication initiation: it is strongly associated with the clinical indication for treatment, carries a high probability of actual statin initiation, and can be identified with sufficient temporal precision in administrative data. This approach should reduce channeling bias, selection bias (e.g., immortal time bias) and residual confounding in future target trial emulations.
- **Broader Significance:** The study provides an empirically derived, high-probability time zero event that can strengthen future target trial emulations using real-world data to assess the safety of commonly used medicines in older adults, a population often underrepresented in randomized trials to obtain regulatory approval.

**PLAIN LANGUAGE SUMMARY:** This study aimed to identify a clear starting point for future research on the safety of atorvastatin in older adults. Using Medicare claims data from 2018–2019, researchers examined more than 1.3 million beneficiaries age 65 and older who had not previously taken statins in the last year. They developed a predictive model to determine which patient characteristics were most strongly linked to starting atorvastatin. The strongest predictors were a recent hospital admission for heart attack (myocardial infarction) or stroke (cerebral infarction). These events were associated with a much higher chance of promptly receiving atorvastatin, which aligns with American College of Cardiology and American Heart Association guidelines recommending statin therapy soon after such events for secondary prevention. By using hospital discharge after these acute events as the starting point for future studies, researchers can create comparisons that reduce bias and allow more reliable estimates of atorvastatin’s effects on potential harms in this vulnerable elderly population.

## INTRODUCTION

Pharmacoepidemiologic studies seeking to emulate target trials using real-world data must emulate not only randomization but also eligibility criteria, treatment assignment, and follow-up rules of the hypothetical trial.^1–3^ A critical design decision is the choice of time zero or the time zero event that anchors eligibility assessment, exposure classification, and follow-up initiation.^1,2,4^ When the exposure of interest corresponds to medication initiation, we postulate that time zero should be: 1) strongly associated with the clinical indication for treatment; 2) associated with a reasonably high probability of actual initiation; and 3) measurable with sufficient temporal precision in administrative data. Failure on any of these three criteria, including insufficient variability in the probability of treatment initiation after time zero, may introduce channeling bias, thereby undermining clinical equipoise and potentially leading to unmeasurable residual confounding.

In pharmacoepidemiologic research using observational data (e.g., health insurance claims data), where laboratory values and detailed clinical notes are typically unavailable, investigators cannot assume that clinical events suggested by guidelines or clinical knowledge will necessarily satisfy the proposed criteria for a valid time zero. A diagnosis or procedure aligned with a therapeutic indication may, for instance, be followed by actual treatment initiation in only a subset of patients. Individuals may decline therapy for varied and often unmeasured reasons, including preferences for nonpharmacologic management, concerns about adverse events, or general reluctance to initiate medication. Because of this heterogeneity, anchoring time zero to such an event may include many individuals who never start treatment, thereby violating the requirement of high probability of initiation. Moreover, those who do initiate treatment may differ in important, often unmeasured ways from those who do not, introducing channeling bias and potentially residual confounding. In such settings, empirical evaluation of candidate time zero events through predictive modeling in the source data provides a systematic means to assess whether a candidate event adequately satisfies the proposed conditions.

Using Medicare claims data, we illustrate how empirical predictive modeling can augment clinical knowledge and guideline recommendations to identify suitable time zero events. For statins such as atorvastatin, several candidate time zero events recorded in administrative data warrant consideration. These include a diagnosis of hyperlipidemia, events related to the primary prevention of atherosclerotic cardiovascular disease, and acute events such as myocardial infarction or ischemic stroke that prompt guideline-directed statin therapy for secondary prevention. We exemplify our eight-step procedure in this context to illustrate the potential value of an empirical approach even when clinical knowledge and guidelines exist.

## METHODS

### Study Design, Data Source and Study Population

We conducted a retrospective cohort study using Medicare fee-for-service claims from Parts A (inpatient), B (outpatient), and D (prescription drugs) during the study period January 1, 2018, through December 31, 2019. The study population consisted of Medicare beneficiaries who, on the study period start date, were age ≥ 65 years, had at least 12 months of continuous enrollment in Parts A, B, and D, and had no recorded statin dispensing in the prior 12 months.

### Exposure Classification

For all patients in the study population, the index date was the date of the first pharmacy dispensing claim for a newly initiated outpatient medication during the study period. Patients were classified as atorvastatin initiators if atorvastatin was dispensed on that date, and as non-atorvastatin initiators otherwise. To balance computational feasibility with statistical precision, we retained a 10% random sample of non-atorvastatin initiators. Patients who did not initiate a new medication during the study period were not included in the study cohort.

### Candidate Predictor Variables

Candidate predictor variables were ascertained during the 6 months immediately preceding the index date (inclusive of the index date unless otherwise specified). The rationale for using a 6-month vs a 12 month pre-index ascertainment period was to require a closer temporal relationship between incident medication initiation and patient level factors documented during routine care (e.g., inpatient admission and pharmacotherapy utilization). Baseline candidate predictor variables were dichotomized to facilitate clinical interpretation and model stability. Demographic characteristics, including age, sex, race/ethnicity, dual Medicare/Medicaid eligibility status, disability status, and geographic region, were obtained from the Medicare Beneficiary Summary File.

Comorbidities were ascertained from both inpatient and outpatient claims using International Classification of Diseases, Tenth Revision, Clinical Modification (ICD-10-CM) codes. These codes were grouped into 552 categories using the Healthcare Cost and Utilization Project Clinical Classifications Software Refined (CCSR) for ICD-10-CM diagnoses.^5^ Separate binary candidate predictor variables were created for each CCSR category from inpatient diagnoses (restricted to the first and second position) and from outpatient diagnoses (any position). The Charlson Comorbidity Index was calculated as a continuous score, then dichotomized at the 75th percentile.

During the 6-month baseline period, healthcare resource utilization was measured by emergency department visits and inpatient admissions, expressed as binary indicators. Total medical costs were summed across all service types and dichotomized at the 75th percentile.

Baseline pharmacotherapy utilization was ascertained from Medicare Part D claims data using generic names and Generic Code Drug Formulary (GCDF) codes. These data were linked to the First Data Bank (FDB) medication classification system and the hierarchical coding structure of the American Hospital Formulary Service (AHFS). Therapeutic categories judged non-relevant to the study objectives were excluded according to prespecified criteria, leaving 20 major pharmacologic-therapeutic classes (using the first two digits of the AHFS code) and 397 subclasses (using the first six digits of the AHFS code). The resulting classification system produced binary indicator variables tied to generic drug names and formulation codes, yielding a total of 417 binary medication-related candidate predictor variables that were applied to Part D claims to classify pharmacotherapy exposure in the 6 months before the index date (excluding the index date itself).

### Statistical Analysis

We applied a prespecified eight-step procedure to identify baseline characteristics associated with incident atorvastatin initiation among statin-naïve Medicare beneficiaries age 65 years or older, relative to those who newly initiated a different medication. Importantly, our goal was not to estimate a causal effect of atorvastatin initiation or to balance baseline characteristics between groups, as would be appropriate in comparative effectiveness or safety research. In predictive modeling for this purpose, controlling for confounding or balancing baseline characteristics is unnecessary and would undermine the goal, as such steps would obscure the very differences we aimed to detect. Instead, we sought to empirically characterize the baseline characteristics that distinguish statin-naïve patients initiating atorvastatin from patients initiating other medications, thereby identifying suitable time-zero events for future target trial emulations. Additionally, this procedure provides guidance on other variables that may not be suitable as time-zero events but that may be used to balance initiators and non-initiators in comparative research.

Details of the structured eight-step candidate predictor variable selection procedure are presented in **Table 1**. In brief, candidate predictors first underwent data management steps consisting of exclusion of continuous variables with any missing values, dichotomization of the remaining continuous variables at prespecified clinically relevant cut points (or at the 75th percentile when no such cut point existed), and removal of binary variables with any cell count below 40. Variables retained after these steps were further screened for imbalance and correlation. We excluded variables with standardized mean differences (SMD) below 0.1, given the large sample size. Among the remaining variables, we assessed collinearity and removed one variable from each pair with an absolute correlation coefficient greater than 0.8, retaining the variable with the stronger univariate association with the exposure variable (atorvastatin vs non-atorvastatin initiator) and giving preference to inpatient diagnoses when associations were similar. All analyses were performed for the primary and secondary cohorts using Stata version 18.0 (College Station, TX).

**Table 1.** Eight-step predictive modeling procedure.

|  |  |
| --- | --- |
| Step 1 | Missingness was assessed for all continuous candidate predictor variables (CPVs). Any CPV with missing data was excluded from further analysis. |
| Step 2 | Continuous CPVs were dichotomized using a clinically relevant cut point when available, or at the 75th percentile of the distribution otherwise. |
| Step 3 | The prevalence of each CPV was examined in relation to atorvastatin initiation. Any CPV for which any cell in the 2×2 table contained fewer than 40 observations was excluded. Odds ratios and z-statistics were generated for all CPVs that survived this step. |
| Step 4 | Because of the very large sample size, standardized mean differences (SMDs) were used for initial screening rather than hypothesis testing. SMDs were calculated for each CPV between atorvastatin initiators and non-initiators. CPVs with an $SMD \geq 0.1$ were retained and ranked in descending order. CPVs with an $SMD < 0.1$ were not evaluated further as potential predictors of atorvastatin initiation. |
| Step 5 | Pairwise Pearson correlations were examined to assess collinearity between CPVs. When the absolute correlation between two CPVs reached or exceeded 0.8, the variable with the stronger association with atorvastatin initiation was retained. Strength of association was quantified as the product of the absolute z-statistic and the odds ratio obtained from univariable logistic regression. This metric incorporates both the magnitude of the effect and its statistical precision. In addition, when resolving collinearity, variables based on inpatient diagnoses were given greater weight (inpatient diagnosis weight = 2; outpatient diagnosis weight = 1). The variable with the lower combined strength of association was excluded. |
| Step 6a | Predictor selection stability was evaluated using Monte Carlo simulation with adaptive LASSO regression for all CPVs retained after Step 5. In each of 1,000 bootstrap replications, a sample of 10,000 observations was drawn with replacement and adaptive LASSO regression was applied. The percentage of replications in which each CPV received a nonzero coefficient was recorded. |
| Step 6b | The Monte Carlo simulation was repeated using bootstrapped stepwise regression for all CPVs retained after Step 5. In each of 1,000 bootstrap replications (n=10,000 with replacement), backward stepwise logistic regression was performed with $p=0.01$ for entry and $p=0.05$ for retention. The percentage of replications in which each CPV was selected was recorded. |
| Step 7 | CPVs were retained and included in the final predictive model if they were selected in at least 80% of the adaptive LASSO replications and at least 80% of the stepwise logistic regression replications. |
| Step 8 | The final model was fitted using multivariable logistic regression. Odds ratios and 95% confidence intervals were obtained with bootstrap standard errors based on 1,000 replications with replacement. Discrimination was assessed with the C-statistic. Calibration was assessed visually by plotting the observed proportion of atorvastatin initiation against the model-predicted probability, grouped into 1,000 quantiles. |

Predictor selection stability was evaluated using two independent Monte Carlo simulation approaches, each consisting of 1,000 bootstrap samples of 10,000 observations drawn with replacement. One approach used adaptive LASSO regression (least absolute shrinkage and selection operator); the other used backward stepwise logistic regression. These two methods are complementary because they relied on fundamentally different mechanisms for variable selection. Adaptive LASSO applies a penalty that shrinks the coefficients of less important candidate predictor variables toward zero and removes them entirely from the model when their contribution is minimal, thereby performing simultaneous variable selection while keeping the model from becoming overly complex. In contrast, backward stepwise logistic regression begins with the full model containing all candidate predictor variables and sequentially removes the least statistically significant variable (i.e., the variable with the largest p-value) one at a time until all remaining variables meet the prespecified significance threshold. This approach is valid for identifying significant, independent predictors of atorvastatin initiation (versus initiation of another medication) because it empirically isolates those baseline characteristics that retain independent associations after mutual adjustment for all other candidates.

Candidate predictor variables selected in at least 80% of bootstrap replications in both the adaptive LASSO and backward stepwise logistic regression approaches comprised the final set of predictors. The final model was fit by multivariable logistic regression, with bootstrap standard errors derived from 1,000 replications. Model discrimination was quantified with the C-statistic. Calibration was examined by plotting observed versus predicted probabilities after grouping observations into 1,000 quantiles of predicted risk.

From the final model, we estimated marginal predicted probabilities of atorvastatin initiation for prespecified patient profiles by fixing the profile-defining covariates at the values of interest and averaging the model-based predictions over the observed empirical distribution of the remaining covariates. This standardization approach yielded valid estimates of the expected probability under each prespecified patient profile by averaging the individual counterfactual predictions obtained by assigning the profile-specific values while retaining observed values for the other covariates, thereby marginalizing properly over the empirical joint distribution without unrealistic assumptions of independence or arbitrary reference values.

To evaluate whether seasonality or temporal trends affected the identification of baseline factors associated with atorvastatin initiation, we conducted a sensitivity analysis restricted to new medication starts occurring on or after July 1, 2018 (i.e., six months after the study period start date).

## RESULTS

The study population included 4,071,007 statin-naïve Medicare beneficiaries age 65 years and older who met the eligibility criteria. The study cohort (N = 510,152) comprised 117,069 incident atorvastatin initiators and 393,083 non-atorvastatin initiators (10% random sample of eligible non-atorvastatin initiators) based on the first pharmacy dispensing claim for a newly initiated medication during the study period. The distribution of index dates was similar for atorvastatin initiators and non-atorvastatin initiators (Supplemental Material **Figure S1**). In both groups, the density of new medication starts was highest immediately after the study period start date and declined progressively thereafter.

We identified 1,532 candidate predictor variables from demographic, healthcare resource utilization, inpatient and outpatient diagnoses, and outpatient medication claims. Through the eight-step variable selection procedure, we sequentially reduced this set by excluding variables with low prevalence (Step 3: 784 retained), weak associations with atorvastatin initiation (Step 4: 113 retained), and high collinearity (Step 5: 110 retained). After evaluating predictor selection stability through Monte Carlo simulation with adaptive LASSO regression (Step 6a: 41 retained) and backward stepwise logistic regression (Step 6b: 19 retained), nineteen candidate predictor variables were retained in the final predictive model. Details of variable selection and attrition across modeling steps and variable types is provided in Supplemental Material **Table S1**.

The strongest predictors of atorvastatin initiation were recent inpatient admissions for cerebral infarction (OR 11.29, 95% CI 10.47-12.17) and acute myocardial infarction (OR 6.56, 95% CI 5.92-7.27). These two variables were the most stable predictors; each selected in nearly all bootstrap replications by both adaptive LASSO and stepwise regression. Female sex, White race and age ≥ 80 years were associated with a lower likelihood of atorvastatin initiation. All nineteen significant, independent predictors of atorvastatin initiation included in the final multivariable model are presented in **Figure 2** and in Supplemental Material **Table S2**. Additional results (e.g., univariate associations and Monte Carlo simulation results) are provided in Supplemental Material **Table S3**.

**Figure 1.**
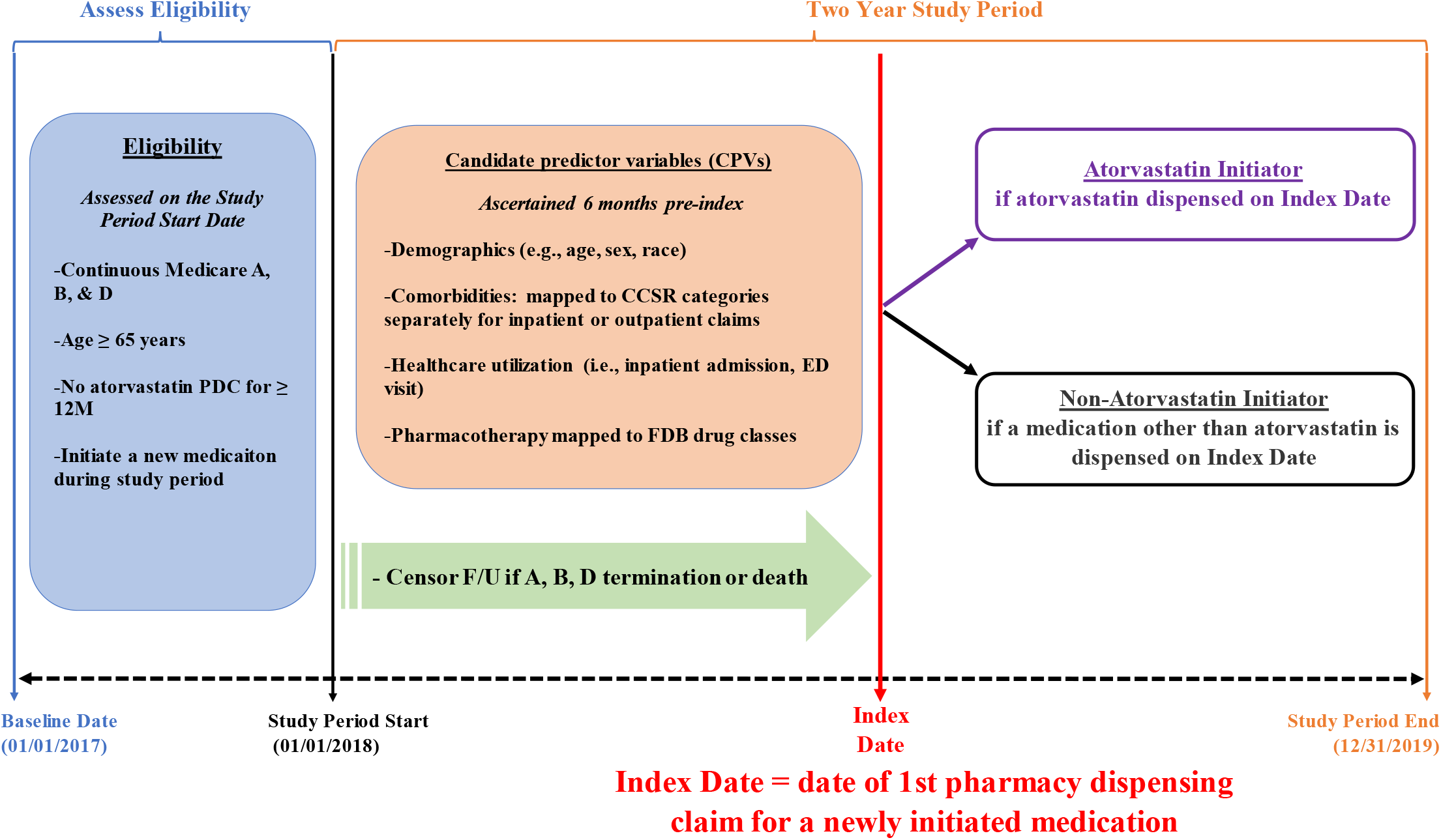
Study Schema. Study design schematic illustrating eligibility assessment, ascertainment of candidate predictor variables (CPVs), and construction of the atorvastatin initiator and non-atorvastatin initiator cohorts during the two-year study period (2018–2019). Eligibility was assessed on the study period start date (January 1, 2018). Candidate predictor variables were ascertained during the 6 months before the index date. The index date was defined as the date of the first pharmacy dispensing claim for a newly initiated medication. Patients were classified as atorvastatin initiators if atorvastatin was dispensed on the index date and as non-atorvastatin initiators otherwise. Follow-up was censored upon termination of Medicare Parts A, B, or D coverage or death. **Abbreviations**: CPVs = candidate predictor variables; PDC = pharmacy dispensing claim; ED = emergency department; FDB = First Databank; F/U = follow-up.

**Figure 2.**
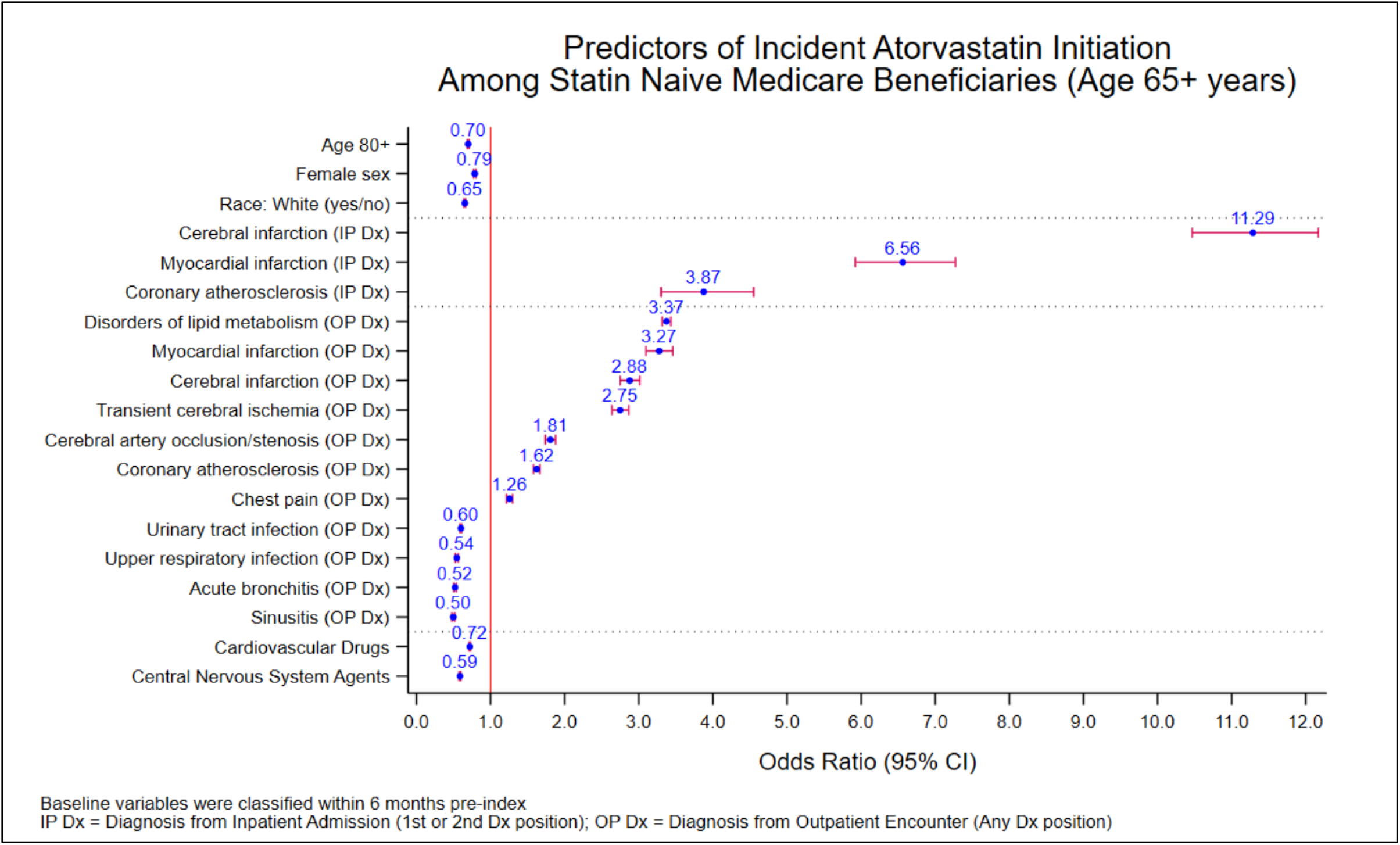
Final Predictive Model.

Model performance diagnostics were assessed to empirically evaluate the final multivariable model’s ability to distinguish individuals who initiated atorvastatin from those who did not (discrimination), and how closely the model’s predicted probabilities of atorvastatin initiation match the observed proportion of individuals who initiated atorvastatin (calibration). The final model achieved a C-statistic of 0.78 with excellent calibration (**Figure 3**).

**Figure 3.**
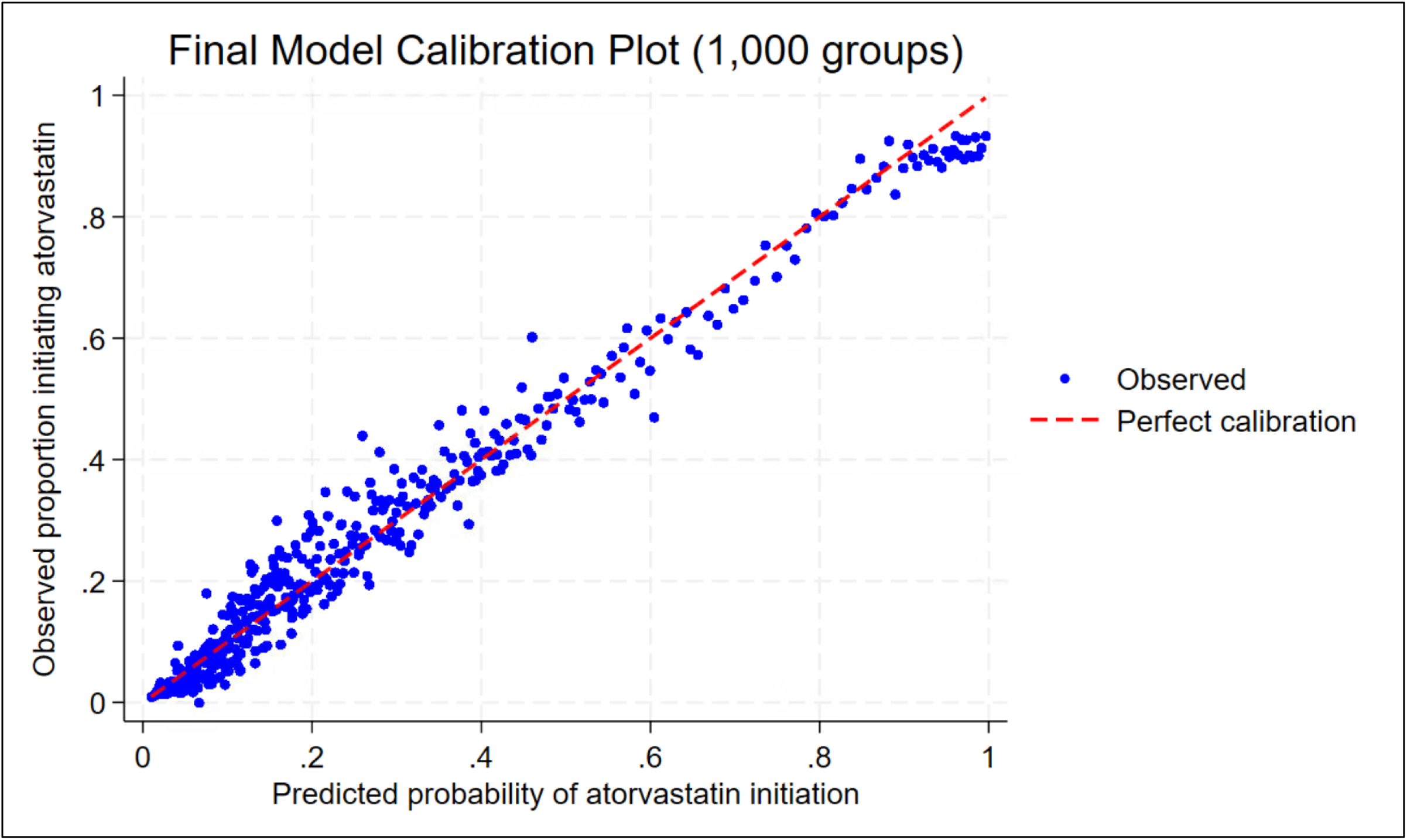
Calibration Plot of Final Predictive Model.

To illustrate the potential utility of empirical variable selection to guide the definition of a time zero event, marginal predicted probabilities from the final model showed that a White male with a recent inpatient cerebral infarction had an 69% probability of atorvastatin initiation (95% CI 68-70). The corresponding probability after inpatient acute myocardial infarction was 59% (95% CI 56-61). In contrast, these probabilities fell to 41% (95% CI 40-42) and 44% (95% CI 43-45), respectively, when the same diagnoses were recorded only in outpatient claims, and to 34% (95% CI 34-35) with an outpatient diagnosis of hyperlipidemia.

The sensitivity analysis restricted to index dates occurring six months after the study period start date (July 1, 2018) included 158,284 Medicare beneficiaries: 42,487 atorvastatin initiators and 115,797 non-atorvastatin initiators (Supplemental Material **Table S4 and S5**). Consistent with the primary analysis, inpatient diagnoses of cerebral infarction (OR 9.66, 95% CI 8.76–10.65) and acute myocardial infarction (OR 4.62, 95% CI 3.88–5.49) remained the strongest predictors of atorvastatin initiation.

## DISCUSSION

We conducted this study to empirically identify measurable baseline factors that strongly predict incident atorvastatin initiation in older, statin naïve Medicare beneficiaries. At the outset, it was unclear whether commonly considered events, such as an outpatient diagnosis of hyperlipidemia or coronary atherosclerosis, would satisfy the three proposed criteria to classify the time zero event for future target trial emulations.

The predictive modelling analyses identified nineteen significant, independent predictors of atorvastatin initiation. Acute myocardial infarction and cerebral infarction recorded during an inpatient admission were the strongest predictors. These two predictors are clinically coherent, directly measurable in Medicare claims data, and satisfy the three key requirements for a valid time zero event when studying medication initiation: strong association with the clinical indication for treatment, reasonably high probability of actual treatment initiation, and measurable with sufficient temporal precision. The predicted probability results illustrated that inpatient admissions for cerebral and myocardial infarction provide a stronger and more specific signal for atorvastatin initiation than outpatient diagnoses of these conditions or an outpatient diagnosis of hyperlipidemia. Results from the sensitivity analysis restricted to index dates six months after the study period start date, were consistent with the primary findings and showed that the identified predictors were independent of seasonal or temporal trends. Accordingly, defining time zero at hospital discharge after these inpatient events should yield more comparable groups at baseline, thereby improving exchangeability and better satisfying the eligibility and treatment assignment criteria for future target trial emulations.

The key findings align closely with current American College of Cardiology and American Heart Association guidelines, which recommend prompt statin therapy for secondary prevention following acute myocardial infarction or ischemic stroke.^6^ Defining the time zero event as the discharge date following these events allows future target trial emulations to synchronize eligibility assessment, treatment assignment, and the start of follow-up at a point of high treatment initiation probability. This approach mitigates channeling bias while providing a reliable, claims-derived time zero event for subsequent trial emulations of atorvastatin safety in elderly Medicare beneficiaries.

Although inpatient diagnoses of acute myocardial infarction and cerebral infarction may serve as central components for defining time zero in future trial emulations for atorvastatin in older statin naïve patients, the prevalence of these predictors requires careful consideration with respect to sample size and statistical precision. Investigators should carefully examine empirical frequency distributions of strong predictors that may be used to define time zero and determine whether they occur with sufficient frequency to support the sample size required for a future trial emulations. When a single predictor proves too infrequent, composite time-zero definitions incorporating multiple predictors (e.g., inpatient diagnosis of cerebral infarction OR myocardial infarction) may be considered to achieve an appropriate balance between methodologic validity and feasibility. In addition, predictors retained in the final model but not used for defining time zero should be addressed analytically, through their inclusion in propensity score models or as covariates in the outcome model, to balance baseline characteristics between initiators and non-initiators and thereby mitigate channeling bias.

In addition to considerations of prevalence, the selection of time zero must also address the proposed requirement that a suitable event has a reasonably high probability of actual treatment initiation. This criterion is inherently nuanced. Preferred events are those that approximate clinical equipoise, the condition under which randomization might be ethically and practically acceptable in an experimental trial. In observational research, investigators should therefore favor time zero events associated with a probability of actual treatment initiation that is substantially greater than zero but well below certainty.

The predictive modeling approach itself requires comment. In a dataset of this size, even trivial differences between atorvastatin initiators and non-initiators attained nominal statistical significance in univariable logistic regression. We therefore relied on standardized mean differences (SMDs) with a threshold of ≥ 0.1, rather than p-values, for initial screening of candidate predictors. This effect-size-based criterion directed attention toward variables with clinically meaningful imbalances rather than associations that were statistically significant merely because of the large sample size. Bootstrap validation, conducted with both adaptive LASSO regression and backward stepwise logistic regression, provided quantitative protection against overfitting and permitted evaluation of the stability of variable selection. The final model, which retained all selected predictors, achieved a C-statistic of 0.78. This level of discrimination is more than adequate for the present purpose of identifying a reliable time zero event, although it would be modest for individual-level prediction.

Additional limitations arise from the data source. Medicare claims data do not include direct laboratory measurements, such as LDL cholesterol levels, or detailed information on lifestyle factors (e.g., smoking status). These unmeasured variables may have served as strong predictors of atorvastatin initiation, and their absence may have reduced the completeness of our predictive model. In addition, we dichotomized continuous variables such as age and the Charlson Comorbidity Index. Although this approach entails some loss of information, it facilitated stable modeling and Monte Carlo simulations for predictor selection while producing clinically relevant thresholds that are straightforward to interpret and implement in future target trial emulations. External validation in an independent database has not yet been performed; without it, the stability and transportability of the identified predictors remain uncertain. Finally, these results derive from statin-naïve fee-for-service Medicare beneficiaries age 65 years and older and may not generalize to other populations, such as those enrolled in Medicare Advantage plans or younger commercially insured individuals.

## Conclusion

The selection of a valid time zero event is essential to the internal validity of target trial emulations conducted with real-world data. In this predictive modeling study, acute myocardial infarction and cerebral infarction recorded during an inpatient admission emerged as the two strongest independent predictors of incident atorvastatin initiation among statin-naïve Medicare beneficiaries age 65 years and older. These acute cardio- and cerebrovascular events were associated with markedly elevated odds of atorvastatin initiation, consistent with guideline-directed secondary prevention. By anchoring the time zero event to hospital discharge following these diagnoses and accounting for the other predictors in the model, future target trial emulations in drug safety research may better synchronize eligibility, treatment assignment, and follow-up at a point of high treatment probability, thereby minimizing channeling bias, selection bias and potentially residual (unmeasured) confounding.

## ETHICS STATEMENT

The research program pertaining to this study, titled “Pharmacovigilance 2.0: Elderly, Disabled, and Disadvantage Populations”, was reviewed and approved by the Institutional Review Board (IRB) at Rutgers University Human Research Protection Program (HRPP) on March 12, 2024. The IRB classified the research as exempt under categories 4(ii) and 4(iii) of the U.S. Code of Federal Regulations, Title 45, Part 46 (45 CFR 46), determining it poses minimal risk. Category 4(ii) applies to research using existing, anonymized data or records where participants cannot be identified, while category 4(iii) covers benign behavioral interventions or interactions with adults where identifiable information is protected, and disclosure poses no significant risk. The principal investigator, Christopher G. Rowan PhD, conducted the study with approval under eIRB protocol #Pro2024000224, supported by the Veritas Research Foundation.

## DATA AVAILABILITY STATEMENT

This study was completed in part by data resources provided by the Institute for Health Data Core at Rutgers University, available at: http://www.ifhcore.rutgers.edu. The data supporting the research initiative, titled *Pharmacovigilance 2*.*0: Elderly, Disabled, and Disadvantage Populations* and conducted under the principal investigator Christopher Rowan, were obtained from the Centers for Medicare & Medicaid Services (CMS) under Data Use Agreement number RSCH-2024-70249. These data contain protected health information subject to the Privacy Act and HIPAA regulations. Due to privacy, confidentiality, and contractual restrictions imposed by CMS, the datasets are not publicly available and cannot be shared by the authors. Qualified researchers interested in accessing similar data may apply independently through the Research Data Assistance Center (ResDAC) at https://resdac.org/ and execute their own Data Use Agreement with CMS.

## ACKNOWLEDGEMENTS

We express our sincere gratitude to Jesse Berlin, ScD, Dan Horton, MD, MSCE, and Tobi Gerhard, BSPharm, PhD, FISPE, for their invaluable scientific and methodologic support. We are deeply thankful to Mathew Iozzio, Haoqian Chen PhD, and the Center for Pharmacoepidemiology Treatment Science at Rutgers University for their partnership on Pharmacovigilance 2.0. We also appreciate the Center for Medicare and Medicaid Services (CMS) for providing data access and support for this initiative. Heartfelt thanks go to the Veritas Research Foundation Board of Directors - Scott C. Durbin, MPH, Mathew Allard, MBA, Bradley J. Buecker, MFA, Gregory Lobdell, MA, and LeeAnn Hard, CPA—for their steadfast trust and support, and to U.S. Medicare beneficiaries for trusting the Veritas Research Foundation to conduct independent drug safety research on their behalf.

