## Supplemental Tables S1-S5 and Figures S1-S2 for "Can Predictive Modeling Inform the Selection of Time Zero for Target Trial Emulations? An Empirical Study of Atorvastatin Initiation in Medicare Beneficiaries"

**Supplemental material table legend**

- **Table S1**
  - Candidate Predictor Variable Selection and Attrition Across Modeling Steps
- **Table S2**
  - Final Predictive Model
- **Table S3**
  - Predictive Modeling and Univariate Results
- **Table S4**
  - Sensitivity Analysis - Final Predictive Model
- **Table S5**
  - Sensitivity Analysis - Predictive Modeling and Univariate Results

**Supplemental material figure legend**

- **Figure S1**
  - Distribution of Index Dates for Atorvastatin Initiators and Non- Atorvastatin Initiators
- **Figure S2**
  - Sensitivity Analysis - Distribution of Index Dates for Atorvastatin Initiators and Non- Atorvastatin Initiators

**Table S1 Predictive Modeling Procedure: Candidate Predictor Variable Selection and Attrition Across Modeling Steps**

| Step | Description | # CPVs Retained After Executing Each Step | | | | | |
|---|---|---|---|---|---|---|---|
| | | Demo | HRU | IP Dx | OP Dx | OP Rx | Total |
| 1 | Assessed missingness; excluded vars w/ missing data | 9 | 2 | 552 | 552 | 417 | **1532** |
| 2 | Dichotomized continuous CPVs (clinical cut or 75th) | 9 | 2 | 552 | 552 | 417 | **1532** |
| 3 | Excluded rare CPVs (<40 events) | 9 | 2 | 158 | 421 | 194 | **784** |
| 4 | Calculated SMDs; retained CPVs with SMD ≥ 0.1 | 5 | 1 | 11 | 63 | 33 | **113** |
| 5 | Removed highly correlated CPVs (\|r\| ≥ 0.8); kept stronger | 5 | 0 | 11 | 61 | 32 | **109** |
| 6a | Monte Carlo simulation: Adaptive LASSO regression | 3 | 0 | 4 | 25 | 9 | **41** |
| 6b | Monte Carlo simulation: Stepwise logistic regression | 3 | 0 | 3 | 11 | 2 | **19** |
| 7 | Retained CPVs selected in ≥ 80% of both simulations | 3 | 0 | 3 | 11 | 2 | **19** |
| 8 | Fitted final model | 3 | 0 | 3 | 11 | 2 | **19** |

CPV = candidate predictor variable

Demo = demographics; HRU = health resource utilization; IP Dx = inpatient diagnosis (1st or 2nd position); OP Dx = outpatient diagnosis (any position); OP Rx = outpatient prescription medication ascertained using pharmacy dispensing claims (Part D)

**Table S2 Final Predictive Model**

| Predictors of Atorvastatin Initiation | Odds Ratio | LCL | UCL | Std Error | Z |
|---|---|---|---|---|---|
| ***Demographics*** | | | | | |
| Female sex | 0.79 | 0.77 | 0.80 | 0.008 | -22.48 |
| Age 80+ | 0.70 | 0.69 | 0.71 | 0.006 | -41.26 |
| Race: White (yes/no) | 0.65 | 0.64 | 0.66 | 0.006 | -47.74 |
| ***Inpatient Diagnosis*** | | | | | |
| Cerebral infarction | 11.29 | 10.47 | 12.17 | 0.434 | 62.98 |
| Myocardial infarction | 6.56 | 5.92 | 7.27 | 0.344 | 35.93 |
| Coronary atherosclerosis | 3.87 | 3.30 | 4.55 | 0.318 | 16.53 |
| ***Outpatient Diagnosis*** | | | | | |
| Disorders of lipid metabolism | 3.37 | 3.31 | 3.43 | 0.030 | 135.42 |
| Myocardial infarction | 3.27 | 3.10 | 3.46 | 0.093 | 41.91 |
| Cerebral infarction | 2.88 | 2.75 | 3.01 | 0.069 | 44.36 |
| Transient cerebral ischemia | 2.75 | 2.64 | 2.86 | 0.057 | 48.58 |
| Cerebral artery occlusion/stenosis | 1.81 | 1.74 | 1.88 | 0.036 | 30.02 |
| Coronary atherosclerosis | 1.62 | 1.58 | 1.67 | 0.022 | 35.16 |
| Chest pain | 1.26 | 1.21 | 1.30 | 0.021 | 13.52 |
| Urinary tract infections | 0.60 | 0.59 | 0.61 | 0.005 | -62.31 |
| Other specified upper respiratory infections | 0.54 | 0.52 | 0.56 | 0.010 | -31.61 |
| Acute bronchitis | 0.52 | 0.50 | 0.54 | 0.008 | -41.8 |
| Sinusitis | 0.50 | 0.48 | 0.51 | 0.009 | -37.28 |
| ***Pharmacotherapy*** | | | | | |
| Cardiovascular Drugs | 0.72 | 0.71 | 0.73 | 0.005 | -48.62 |
| Central Nervous System Agents | 0.59 | 0.58 | 0.60 | 0.005 | -58.21 |

**Table S3 Predictive Modeling and Univariate Results**

| CCSR | Description | Type | LASSO | SWR | SMD | OR | Z |
|---|---|---|---|---|---|---|---|
| CIR020 | Cerebral infarction | IP Dx | 1000 | 1000 | 0.37 | 37.19 | 96.0 |
| END010 | Disorders of lipid metabolism | OP Dx | 1000 | 1000 | 0.60 | 3.29 | 170.0 |
| CIR011 | Coronary atherosclerosis | OP Dx | 1000 | 1000 | 0.38 | 2.75 | 120.0 |
| CIR009 | Acute myocardial infarction | IP Dx | 1000 | 999 | 0.35 | 25.32 | 98.0 |
| - | Age 80+ | Demo | 999 | 999 | 0.10 | 0.80 | -28.0 |
| CIR020 | Cerebral infarction | OP Dx | 1000 | 997 | 0.42 | 7.61 | 135.0 |
| NVS012 | Transient cerebral ischemia | OP Dx | 999 | 996 | 0.27 | 5.43 | 91.0 |
| CIR023 | Occlusion or stenosis | OP Dx | 999 | 991 | 0.33 | 4.33 | 109.0 |
| RSP006 | Other specified upper respiratory infect | OP Dx | 998 | 979 | 0.17 | 0.48 | -45.0 |
| RSP005 | Acute bronchitis | OP Dx | 998 | 979 | 0.16 | 0.47 | -43.0 |
| RSP001 | Sinusitis | OP Dx | 998 | 979 | 0.18 | 0.44 | -47.0 |
| CIR009 | Acute myocardial infarction | OP Dx | 999 | 977 | 0.37 | 12.45 | 117.0 |
| GEN004 | Urinary tract infections | OP Dx | 993 | 955 | 0.10 | 0.72 | -29.0 |
| - | Cardiovascular Drugs | OP Rx | 998 | 928 | 0.13 | 0.78 | -38.0 |
| - | Central Nervous System Agents | OP Rx | 999 | 915 | 0.28 | 0.55 | -82.0 |
| CIR011 | Coronary atherosclerosis | IP Dx | 995 | 895 | 0.19 | 12.77 | 59.0 |
| CIR012 | Nonspecific chest pain | OP Dx | 981 | 842 | 0.26 | 2.21 | 84.0 |
| - | Race: White (yes/no) | Demo | 951 | 822 | 0.17 | 0.65 | -52.0 |
| - | Female sex | Demo | 983 | 802 | 0.22 | 0.63 | -67.0 |
| MUS006 | Osteoarthritis | OP Dx | 957 | 792 | 0.13 | 0.72 | -39.0 |
| END005 | Diabetes mellitus, Type 2 | OP Dx | 946 | 782 | 0.23 | 1.76 | 72.0 |
| CIR007 | Essential hypertension | OP Dx | 981 | 780 | 0.23 | 1.61 | 68.0 |
| SKN001 | Skin and subcutaneous tissue infections | OP Dx | 939 | 779 | 0.11 | 0.58 | -31.0 |
| - | Gastrointestinal Drugs | OP Rx | 966 | 771 | 0.22 | 0.58 | -63.0 |
| NVS012 | Transient cerebral ischemia | IP Dx | 966 | 728 | 0.12 | 15.25 | 37.0 |
| MUS010 | Musculoskeletal pain, not low back pain | OP Dx | 935 | 727 | 0.16 | 0.70 | -47.0 |
| EYE001 | Cornea and external disease | OP Dx | 925 | 726 | 0.16 | 0.55 | -45.0 |
| - | Biguanides | Rx | 830 | 690 | 0.10 | 1.50 | 32.0 |
| EYE002 | Cataract and other lens disorders | OP Dx | 933 | 665 | 0.13 | 0.70 | -38.0 |
| INJ031 | Allergic reactions | OP Dx | 929 | 639 | 0.17 | 0.56 | -47.0 |
| SKN002 | Other specified inflammatory condition | OP Dx | 861 | 626 | 0.15 | 0.55 | -43.0 |

| | | | | | | | |
|---|---|---|---|---|---|---|---|
| SYM001 | Syncope | OP Dx | 298 | 588 | 0.10 | 1.71 | 33.0 |
| SYM012 | Circulatory signs and symptoms | OP Dx | 489 | 588 | 0.15 | 1.52 | 47.0 |
| SYM017 | Abnormal findings without diagnosis | OP Dx | 386 | 588 | 0.19 | 1.49 | 57.0 |
| SYM010 | Nervous system signs and symptoms | OP Dx | 232 | 588 | 0.13 | 1.38 | 41.0 |
| SYM006 | Abdominal pain and other | OP Dx | 999 | 588 | 0.10 | 0.75 | -29.0 |
| SYM011 | Genitourinary signs and symptoms | OP Dx | 920 | 588 | 0.10 | 0.74 | -28.0 |
| - | Loop Diuretics (24 36) | OP Rx | 905 | 588 | 0.11 | 0.66 | -30.0 |
| SKN007 | Other specified and unspecified skin | OP Dx | 965 | 588 | 0.22 | 0.60 | -63.0 |
| SKN005 | Contact dermatitis | OP Dx | 687 | 588 | 0.11 | 0.54 | -31.0 |
| INF004 | Fungal infections | OP Dx | 852 | 549 | 0.13 | 0.67 | -36.0 |
| - | Anti-infective Agents | OP Rx | 883 | 528 | 0.26 | 0.49 | -73.0 |
| MUS011 | Spondylopathies/spondyloarthropathy | OP Dx | 739 | 524 | 0.13 | 0.71 | -39.0 |
| - | Hormones and Synthetic Substitutes | OP Rx | 912 | 515 | 0.16 | 0.71 | -47.0 |
| - | Eye, Ear, Nose, and Throat Preparations | OP Rx | 884 | 506 | 0.25 | 0.53 | -70.0 |
| CIR019 | Heart failure | IP Dx | 714 | 495 | 0.17 | 3.39 | 58.0 |
| NEO072 | Neoplasms of unspecified nature | OP Dx | 803 | 470 | 0.12 | 0.62 | -35.0 |
| - | Autonomic Drugs | OP Rx | 851 | 467 | 0.11 | 0.78 | -33.0 |
| EYE003 | Glaucoma | OP Dx | 709 | 463 | 0.11 | 0.69 | -32.0 |
| FAC020 | Lifestyle/life management factors | OP Dx | 720 | 443 | 0.11 | 1.97 | 34.0 |
| - | Thyroid Agents | OP Rx | 686 | 410 | 0.17 | 0.62 | -48.0 |
| CIR024 | Other and ill-defined cerebrovascular | OP Dx | 694 | 392 | 0.25 | 4.01 | 83.0 |
| EYE005 | Retinal and vitreous conditions | OP Dx | 746 | 379 | 0.12 | 0.72 | -34.0 |
| CIR015 | Other and ill-defined heart disease | OP Dx | 605 | 365 | 0.20 | 2.34 | 65.0 |
| MBD024 | Tobacco-related disorders | OP Dx | 679 | 341 | 0.12 | 1.80 | 39.0 |
| EAR006 | Other specified and unspecified disorder | OP Dx | 688 | 327 | 0.10 | 0.66 | -27.0 |
| - | Respiratory Tract Agents | Rx | 608 | 300 | 0.15 | 0.56 | -42.0 |
| NVS020 | Other nervous system disorders | IP Dx | 522 | 288 | 0.12 | 2.79 | 39.0 |
| CIR008 | Hypertension with complications | IP Dx | 600 | 276 | 0.14 | 3.33 | 47.0 |
| CIR017 | Cardiac dysrhythmias | OP Dx | 546 | 248 | 0.11 | 1.34 | 34.0 |
| - | Benzodiazepines | OP Rx | 466 | 246 | 0.15 | 0.54 | -43.0 |
| MUS038 | Low back pain | OP Dx | 554 | 241 | 0.14 | 0.69 | -40.0 |
| - | Race: Hispanic (yes/no) | Demo | 564 | 239 | 0.10 | 1.53 | 30.0 |
| MUS013 | Osteoporosis | OP Dx | 572 | 237 | 0.10 | 0.71 | -28.0 |

| | | | | | | | |
|---|---|---|---|---|---|---|---|
| NEO073 | Benign neoplasms | OP Dx | 582 | 225 | 0.12 | 0.70 | -34.0 |
| MUS025 | Other specified connective tissue | OP Dx | 496 | 219 | 0.10 | 0.70 | -29.0 |
| CIR003 | Nonrheumatic and unspecified valve | OP Dx | 529 | 206 | 0.20 | 1.96 | 63.0 |
| END002 | Diabetes mellitus without complication | OP Dx | 173 | 200 | 0.20 | 1.68 | 62.0 |
| GEN023 | Menopausal disorders | OP Dx | 547 | 192 | 0.10 | 0.55 | -27.0 |
| EYE008 | Oculofacial plastics and orbital | OP Dx | 438 | 188 | 0.13 | 0.62 | -37.0 |
| MUS012 | Biomechanical lesions | OP Dx | 461 | 187 | 0.10 | 0.65 | -27.0 |
| - | Race: Black (yes/no) | Demo | 314 | 171 | 0.11 | 1.52 | 34.0 |
| - | Reversible Cox 1/Cox 2 Inhibitors | OP Rx | 362 | 170 | 0.11 | 0.67 | -31.0 |
| - | Proton Pump Inhibitors | OP Rx | 154 | 169 | 0.15 | 0.65 | -42.0 |
| - | Opioid Agonists (28 08) | OP Rx | 355 | 165 | 0.15 | 0.55 | -42.0 |
| CIR025 | Sequela of cerebral infarction | IP Dx | 294 | 161 | 0.21 | 23.80 | 60.0 |
| - | Corticosteroids (Eent) | OP Rx | 168 | 137 | 0.16 | 0.57 | -46.0 |
| GEN002 | Acute and unspecified renal failure | OP Dx | 356 | 131 | 0.13 | 2.00 | 43.0 |
| RSP012 | Respiratory failure; insufficiency | OP Dx | 306 | 131 | 0.11 | 1.91 | 35.0 |
| - | Adrenals | OP Rx | 193 | 130 | 0.17 | 0.55 | -46.0 |
| CIR005 | Myocarditis and cardiomyopathy | OP Dx | 403 | 124 | 0.15 | 2.26 | 48.0 |
| END003 | Diabetes mellitus with complication | OP Dx | 273 | 123 | 0.17 | 1.70 | 54.0 |
| SYM010 | Nervous system signs and symptoms | IP Dx | 392 | 120 | 0.10 | 5.90 | 34.0 |
| NVS008 | Paralysis (other than cerebral palsy) | OP Dx | 306 | 120 | 0.19 | 5.33 | 65.0 |
| - | Benzodiazepines (Anticonvulsants) | OP Rx | 283 | 120 | 0.13 | 0.53 | -36.0 |
| - | Leukotriene Modifiers | OP Rx | 255 | 119 | 0.10 | 0.50 | -28.0 |
| - | Antiulcer Agents And Acid Suppressant | OP Rx | 135 | 104 | 0.11 | 0.58 | -32.0 |
| CIR001 | Chronic rheumatic heart disease | OP Dx | 341 | 101 | 0.12 | 2.09 | 38.0 |
| - | Antibacterials (52 04) | OP Rx | 161 | 99 | 0.17 | 0.51 | -47.0 |
| CIR021 | Acute hemorrhagic cerebrovascular | OP Dx | 358 | 97 | 0.10 | 3.22 | 32.0 |
| - | Estrogens | OP Rx | 255 | 96 | 0.10 | 0.44 | -26.0 |
| NVS020 | Other nervous system disorders | OP Dx | 264 | 94 | 0.16 | 1.95 | 50.0 |
| CIR017 | Cardiac dysrhythmias | IP Dx | 370 | 93 | 0.10 | 2.52 | 34.0 |
| NVS008 | Paralysis (other than cerebral palsy) | IP Dx | 351 | 88 | 0.19 | 21.24 | 54.0 |
| CIR016 | Conduction disorders | OP Dx | 270 | 84 | 0.11 | 1.52 | 34.0 |
| - | Potassium Competitive Acid Blockers | OP Rx | 152 | 83 | 0.10 | 0.59 | -28.0 |
| - | Gaba Mediated Anticonvulsants | OP Rx | 206 | 81 | 0.11 | 0.64 | -32.0 |

| | | | | | | | |
|---|---|---|---|---|---|---|---|
| CIR025 | Sequela of cerebral infarction | OP Dx | 273 | 77 | 0.23 | 4.70 | 78.0 |
| CIR019 | Heart failure | OP Dx | 237 | 77 | 0.15 | 1.65 | 46.0 |
| - | Serotonin Modulators | OP Rx | 224 | 76 | 0.11 | 0.56 | -30.0 |
| CIR008 | Hypertension with complication | OP Dx | 286 | 70 | 0.16 | 1.62 | 49.0 |
| - | Antimalarials | OP Rx | 206 | 67 | 0.11 | 0.51 | -31.0 |
| - | Skin and Mucous Membrane Agents | OP Rx | 162 | 66 | 0.11 | 0.56 | -31.0 |
| - | Corticosteroids | OP Rx | 108 | 61 | 0.11 | 0.57 | -30.0 |
| RSP011 | Pleurisy, pleural effusion | OP Dx | 267 | 54 | 0.14 | 1.84 | 44.0 |
| - | Selective Serotonin Reuptake Inhibitors | OP Rx | 290 | 0 | 0.12 | 0.66 | -35.0 |
| - | IOP and Antiglaucoma Agents | OP Rx | 352 | 0 | 0.10 | 0.63 | -28.0 |
| - | Other Macrolide Antibiotics | OP Rx | 226 | 0 | 0.11 | 0.56 | -32.0 |
| - | 1St Generation Cephalosporin Antibiotics | OP Rx | 189 | 0 | 0.10 | 0.53 | -28.0 |

Abbreviations: CCSR: Clinical Classifications Software Refined (standardized category codes for clinical conditions, diagnoses, and services); IP Dx: Inpatient diagnosis (1st or 2nd diagnosis position on inpatient claim); OP Dx: Outpatient diagnosis (any diagnosis position on claim); Demo: Demographic variable; OP Rx: Outpatient prescription medication ascertained using pharmacy dispensing claims (Part D); LASSO: Number of times the variable was selected (non-zero coefficient) across 1,000 adaptive LASSO bootstrap replications; SWR: Number of times the variable was selected across 1,000 bootstrapped stepwise logistic regression replications; SMD: Standardized mean difference (between groups); OR: Odds ratio (from univariate analysis); Z: Z-statistic (from univariate analysis)

### Table S4 Sensitivity Analysis - Final Predictive Model

| Predictors of Atorvastatin Initiation | Type | Odds Ratio | LCL | UCL | Std Error | Z |
|---|---|---|---|---|---|---|
| ***Demographics*** | | | | | | |
| Age 80+ | Demographic | 0.64 | 0.62 | 0.66 | 0.01 | -25.65 |
| Female sex | Demographic | 0.79 | 0.77 | 0.82 | 0.01 | -13.83 |
| White Race | Demographic | 0.77 | 0.75 | 0.79 | 0.01 | -18.68 |
| Dual Medicare/Medicaid | Demographic | 1.32 | 1.27 | 1.36 | 0.02 | 15.67 |
| ***Inpatient Diagnosis*** | | | | | | |
| Cerebral infarction | CIR020 | 9.66 | 8.76 | 10.65 | 0.48 | 45.73 |
| Acute myocardial infarction | CIR009 | 4.62 | 3.88 | 5.49 | 0.41 | 17.36 |
| Coronary atherosclerosis and other heart disease | CIR011 | 4.05 | 3.11 | 5.29 | 0.55 | 10.30 |
| ***Outpatient Diagnosis*** | | | | | | |
| Acute myocardial infarction | CIR009 | 4.69 | 4.28 | 5.15 | 0.22 | 32.49 |
| Disorders of lipid metabolism | END010 | 3.50 | 3.40 | 3.60 | 0.05 | 88.69 |
| Transient cerebral ischemia | NVS012 | 3.46 | 3.22 | 3.72 | 0.13 | 33.47 |
| Cerebral infarction | CIR020 | 3.40 | 3.14 | 3.68 | 0.14 | 30.55 |
| Occlusion of cerebral arteries without infarction | CIR023 | 2.10 | 2.01 | 2.19 | 0.05 | 33.78 |
| Coronary atherosclerosis and other heart disease | CIR011 | 1.70 | 1.64 | 1.77 | 0.03 | 26.82 |
| Nonspecific chest pain | CIR012 | 1.42 | 1.36 | 1.48 | 0.03 | 17.03 |
| Diabetes mellitus, Type 2 | END005 | 1.30 | 1.23 | 1.38 | 0.04 | 9.18 |
| Musculoskeletal pain, not low back pain | MUS010 | 0.70 | 0.69 | 0.72 | 0.01 | -30.91 |
| Cataract and other lens disorders | EYE002 | 0.67 | 0.64 | 0.70 | 0.01 | -19.39 |
| Other specified and unspecified skin disorders | SKN007 | 0.66 | 0.64 | 0.68 | 0.01 | -23.35 |
| Allergic reactions | INJ031 | 0.62 | 0.58 | 0.66 | 0.02 | -14.32 |
| Other specified upper respiratory infections | RSP006 | 0.54 | 0.48 | 0.60 | 0.03 | -10.78 |
| Acute bronchitis | RSP005 | 0.45 | 0.41 | 0.48 | 0.02 | -20.80 |
| Sinusitis | RSP001 | 0.43 | 0.40 | 0.45 | 0.01 | -25.41 |
| ***Pharmacotherapy*** | | | | | | |
| Biguanides | Rx | 1.29 | 1.20 | 1.39 | 0.05 | 6.54 |
| Cardiovascular Drug | Rx | 0.72 | 0.71 | 0.74 | 0.01 | -29.00 |
| Central Nervous System Agent | Rx | 0.68 | 0.65 | 0.70 | 0.01 | -19.79 |
| **Constant** | - | 0.33 | 0.31 | 0.34 | 0.01 | -49.10 |

**Table S5 Sensitivity Analysis Restricted to Index Dates ≥ July 1, 2018 - Predictive Modeling and Univariate Results**

| CCSR | Description | Type | LASSO | SWLR | SMD | OR | Z |
|---|---|---|---|---|---|---|---|
| CIR020 | Cerebral infarction | IP Dx | 100 | 100 | 0.4 | 42.15 | 56 |
| CIR009 | Acute myocardial infarction | OP Dx | 100 | 100 | 0.41 | 19.02 | 68 |
| CIR020 | Cerebral infarction | OP Dx | 100 | 100 | 0.47 | 11.06 | 83 |
| NVS012 | Transient cerebral ischemia | OP Dx | 100 | 100 | 0.3 | 7.6 | 56 |
| CIR011 | Coronary atherosclerosis and other heart | OP Dx | 100 | 100 | 0.44 | 3.62 | 81 |
| END010 | Disorders of lipid metabolism | OP Dx | 100 | 100 | 0.62 | 3.36 | 102 |
| Demographic | Age 80+ | Demo | 100 | 100 | 0.1 | 0.8 | -17 |
| Rx | Cardiovascular Drugs | OP Rx | 100 | 100 | 0.11 | 0.79 | -20 |
| MUS010 | Musculoskeletal pain, not low back pain | OP Dx | 100 | 100 | 0.11 | 0.77 | -19 |
| RSP001 | Sinusitis | OP Dx | 100 | 100 | 0.13 | 0.42 | -21 |
| CIR009 | Acute myocardial infarction | IP Dx | 100 | 99 | 0.38 | 29.48 | 57 |
| CIR023 | Occlusion or stenosis of precerebral or | OP Dx | 100 | 99 | 0.38 | 6 | 70 |
| SKN007 | Other specified and unspecified skin dis | OP Dx | 100 | 99 | 0.21 | 0.59 | -36 |
| RSP005 | Acute bronchitis | OP Dx | 100 | 98 | 0.12 | 0.43 | -19 |
| Demographic | Female sex | Demo | 100 | 97 | 0.21 | 0.66 | -37 |
| CIR012 | Nonspecific chest pain | OP Dx | 100 | 94 | 0.34 | 3.12 | 63 |
| Rx | Central Nervous System Agents | OP Rx | 100 | 94 | 0.2 | 0.61 | -34 |
| RSP006 | Other specified upper respiratory infect | OP Dx | 100 | 91 | 0.12 | 0.49 | -20 |
| CIR011 | Coronary atherosclerosis and other heart | IP Dx | 99 | 90 | 0.2 | 17.57 | 34 |
| END005 | Diabetes mellitus, Type 2 | OP Dx | 96 | 90 | 0.26 | 1.97 | 47 |
| INJ031 | Allergic reactions | OP Dx | 100 | 90 | 0.14 | 0.55 | -24 |
| EYE002 | Cataract and other lens disorders | OP Dx | 99 | 87 | 0.16 | 0.65 | -26 |
| Rx | Biguanides | OP Rx | 82 | 84 | 0.1 | 1.59 | 18 |
| Demographic | Dual Medicare/Medicaid | Demo | 100 | 83 | 0.13 | 1.44 | 24 |
| Demographic | Race: White (yes/no) | Demo | 96 | 82 | 0.17 | 0.65 | -30 |
| CIR007 | Essential hypertension | OP Dx | 100 | 79 | 0.29 | 1.8 | 50 |
| Rx | Gastrointestinal Drugs | OP Rx | 92 | 79 | 0.15 | 0.62 | -25 |
| MUS038 | Low back pain | OP Dx | 97 | 74 | 0.1 | 0.73 | -18 |
| Rx | Eye, Ear, Nose, and Throat Preparations | OP Rx | 83 | 67 | 0.16 | 0.56 | -27 |
| EYE005 | Retinal and vitreous conditions | OP Dx | 86 | 62 | 0.14 | 0.67 | -23 |
| SKN002 | Other specified inflammatory condition o | OP Dx | 94 | 62 | 0.14 | 0.54 | -22 |

| | | | | | | | |
|---|---|---|---|---|---|---|---|
| CIR019 | Heart failure | IP Dx | 86 | 61 | 0.21 | 5.45 | 40 |
| CIR024 | Other and ill-defined cerebrovascular di | OP Dx | 73 | 60 | 0.29 | 5.91 | 55 |
| EYE001 | Cornea and external disease | OP Dx | 92 | 60 | 0.15 | 0.53 | -25 |
| NEO072 | Neoplasms of unspecified nature or | OP Dx | 92 | 58 | 0.13 | 0.58 | -21 |
| NVS012 | Transient cerebral ischemia | IP Dx | 95 | 56 | 0.12 | 20.17 | 20 |
| SYM007 | Malaise and fatigue | OP Dx | 91 | 55 | 0.16 | 1.56 | 29 |
| CIR015 | Other and ill-defined heart disease | OP Dx | 72 | 54 | 0.26 | 3.49 | 50 |
| Rx | Anti-infective Agents | OP Rx | 79 | 50 | 0.15 | 0.49 | -23 |
| MBD024 | Tobacco-related disorders | OP Dx | 68 | 47 | 0.16 | 2.19 | 30 |
| Rx | Thyroid Agents | OP Rx | 81 | 47 | 0.15 | 0.62 | -26 |
| CIR008 | Hypertension with complications and | IP Dx | 77 | 46 | 0.18 | 5.56 | 34 |
| SYM013 | Respiratory signs and symptoms | OP Dx | 70 | 46 | 0.13 | 1.41 | 23 |
| Rx | Hormones and Synthetic Substitutes | OP Rx | 89 | 45 | 0.1 | 0.77 | -18 |
| FAC020 | Lifestyle/life management factors | OP Dx | 66 | 42 | 0.13 | 2.48 | 25 |
| HRU | Outpatient office visits 40+: 6M | HRU | 89 | 39 | 0.21 | 1.97 | 39 |
| CIR029 | Aortic; peripheral; and visceral artery | OP Dx | 62 | 37 | 0.11 | 2.23 | 20 |
| EYE003 | Glaucoma | OP Dx | 79 | 35 | 0.12 | 0.65 | -20 |
| END002 | Diabetes mellitus without complication | OP Dx | 17 | 34 | 0.23 | 1.9 | 42 |
| SYM005 | Dysphagia | OP Dx | 72 | 28 | 0.12 | 1.89 | 22 |
| CIR032 | Other specified and unspecified | OP Dx | 51 | 25 | 0.12 | 1.96 | 22 |
| SYM012 | Circulatory signs and symptoms | OP Dx | 67 | 25 | 0.22 | 1.9 | 40 |
| SKN005 | Contact dermatitis | OP Dx | 66 | 25 | 0.11 | 0.51 | -19 |
| CIR003 | Nonrheumatic and unspecified valve | OP Dx | 60 | 23 | 0.25 | 2.47 | 47 |
| CIR017 | Cardiac dysrhythmias | OP Dx | 45 | 23 | 0.18 | 1.67 | 33 |
| HRU | Inpatient Admissions: 6M pre-index | HRU | 96 | 22 | 0.53 | 4.58 | 97 |
| FAC025 | Other specified status | OP Dx | 42 | 22 | 0.15 | 1.35 | 26 |
| END005 | Diabetes mellitus, Type 2 | IP Dx | 54 | 19 | 0.1 | 5.43 | 19 |
| END003 | Diabetes mellitus with complication | OP Dx | 23 | 19 | 0.19 | 1.95 | 35 |
| Demographic | Race: Hispanic (yes/no) | Demo | 56 | 19 | 0.1 | 1.59 | 19 |
| CIR021 | Acute hemorrhagic cerebrovascular | IP Dx | 47 | 18 | 0.1 | 9.61 | 18 |
| RSP012 | Respiratory failure; insufficiency; | IP Dx | 34 | 16 | 0.11 | 3.19 | 22 |
| EYE008 | Oculofacial plastics and orbital | OP Dx | 41 | 16 | 0.12 | 0.62 | -19 |
| Rx | Benzodiazepines | OP Rx | 57 | 16 | 0.1 | 0.57 | -17 |

| | | | | | | | |
|---|---|---|---|---|---|---|---|
| CIR017 | Cardiac dysrhythmias | IP Dx | 45 | 15 | 0.12 | 3.42 | 23 |
| Rx | Proton Pump Inhibitors | OP Rx | 11 | 14 | 0.1 | 0.69 | -17 |
| CIR021 | Acute hemorrhagic cerebrovascular | OP Dx | 30 | 13 | 0.11 | 4.07 | 21 |
| END011 | Fluid and electrolyte disorders | OP Dx | 41 | 13 | 0.17 | 1.89 | 31 |
| CIR026 | Peripheral and visceral vascular disease | OP Dx | 28 | 13 | 0.1 | 1.42 | 18 |
| Rx | Corticosteroids (Eent) | OP Rx | 15 | 13 | 0.1 | 0.59 | -16 |
| SYM003 | Shock | OP Dx | 26 | 12 | 0.1 | 5.34 | 19 |
| GEN003 | Chronic kidney disease | IP Dx | 46 | 12 | 0.11 | 3.85 | 21 |
| NVS020 | Other nervous system disorders (neither | IP Dx | 32 | 12 | 0.14 | 3.7 | 26 |
| SYM010 | Nervous system signs and symptoms | IP Dx | 47 | 11 | 0.12 | 8.9 | 23 |
| Demographic | Race: Black (yes/no) | Demo | 22 | 11 | 0.11 | 1.51 | 19 |
| FAC030 | Personal history of other disease | OP Dx | 37 | 11 | 0.12 | 1.4 | 21 |
| RSP012 | Respiratory failure; insufficiency | OP Dx | 24 | 10 | 0.16 | 3.2 | 30 |
| SYM001 | Syncope | OP Dx | 39 | 10 | 0.14 | 2.17 | 26 |
| CIR016 | Conduction disorders | OP Dx | 30 | 9 | 0.15 | 1.87 | 27 |
| CIR025 | Sequela of cerebral infarction and other | OP Dx | 29 | 8 | 0.28 | 7.37 | 51 |
| CIR001 | Chronic rheumatic heart disease | OP Dx | 36 | 8 | 0.15 | 2.79 | 29 |
| CIR025 | Sequela of cerebral infarction and other | IP Dx | 11 | 7 | 0.23 | 27.87 | 35 |
| NVS008 | Paralysis (other than cerebral palsy) | OP Dx | 25 | 7 | 0.23 | 8.26 | 42 |
| CIR007 | Essential hypertension | IP Dx | 23 | 7 | 0.1 | 3.57 | 19 |
| NVS020 | Other nervous system disorders (neither | OP Dx | 29 | 7 | 0.21 | 2.76 | 39 |
| CIR019 | Heart failure | OP Dx | 20 | 7 | 0.22 | 2.32 | 40 |
| RSP016 | Other specified and unspecified lower re | OP Dx | 34 | 7 | 0.14 | 2.14 | 26 |
| MUS026 | Muscle disorders | OP Dx | 41 | 7 | 0.13 | 1.65 | 25 |
| GEN003 | Chronic kidney disease | OP Dx | 41 | 7 | 0.12 | 1.48 | 21 |
| RSP011 | Pleurisy, pleural effusion and pulmonary | OP Dx | 24 | 6 | 0.21 | 2.93 | 39 |
| CIR005 | Myocarditis and cardiomyopathy | OP Dx | 25 | 6 | 0.18 | 2.93 | 33 |
| SYM015 | General sensation/perception signs and s | OP Dx | 30 | 6 | 0.12 | 1.65 | 21 |
| GEN002 | Acute and unspecified renal failure | IP Dx | 32 | 5 | 0.11 | 3.01 | 20 |
| SYM010 | Nervous system signs and symptoms | OP Dx | 28 | 5 | 0.25 | 1.92 | 46 |
| CIR031 | Hypotension | OP Dx | 26 | 4 | 0.11 | 2.25 | 21 |
| SYM017 | Abnormal findings without diagnosis | OP Dx | 27 | 4 | 0.27 | 1.79 | 48 |
| NVS008 | Paralysis (other than cerebral palsy) | IP Dx | 47 | 3 | 0.21 | 28.34 | 31 |

| | | | | | | | |
|---|---|---|---|---|---|---|---|
| FAC026 | Personal history of nicotine dependence | OP Dx | 25 | 3 | 0.12 | 1.57 | 21 |

Description of column headings: CCSR: Clinical Classifications Software Refined category code and description; Type: Source of the variable (Inpatient, Outpatient, Demographic, Rx, or HRU); LASSO: Number of times the variable was selected (nonzero coefficient) across 1,000 adaptive lasso bootstrap replications; SWLR: Number of times the variable was selected across 1,000 bootstrapped stepwise logistic regression replications (using the swboot command); SMD: Standardized mean difference between atorvastatin initiators and non-initiators; OR: Odds ratio from the final multivariable logistic regression model; Z: z-statistic from the univariate analysis.

**Figure S1 Distribution of Index Dates for Atorvastatin Initiators and Non- Atorvastatin Initiators**

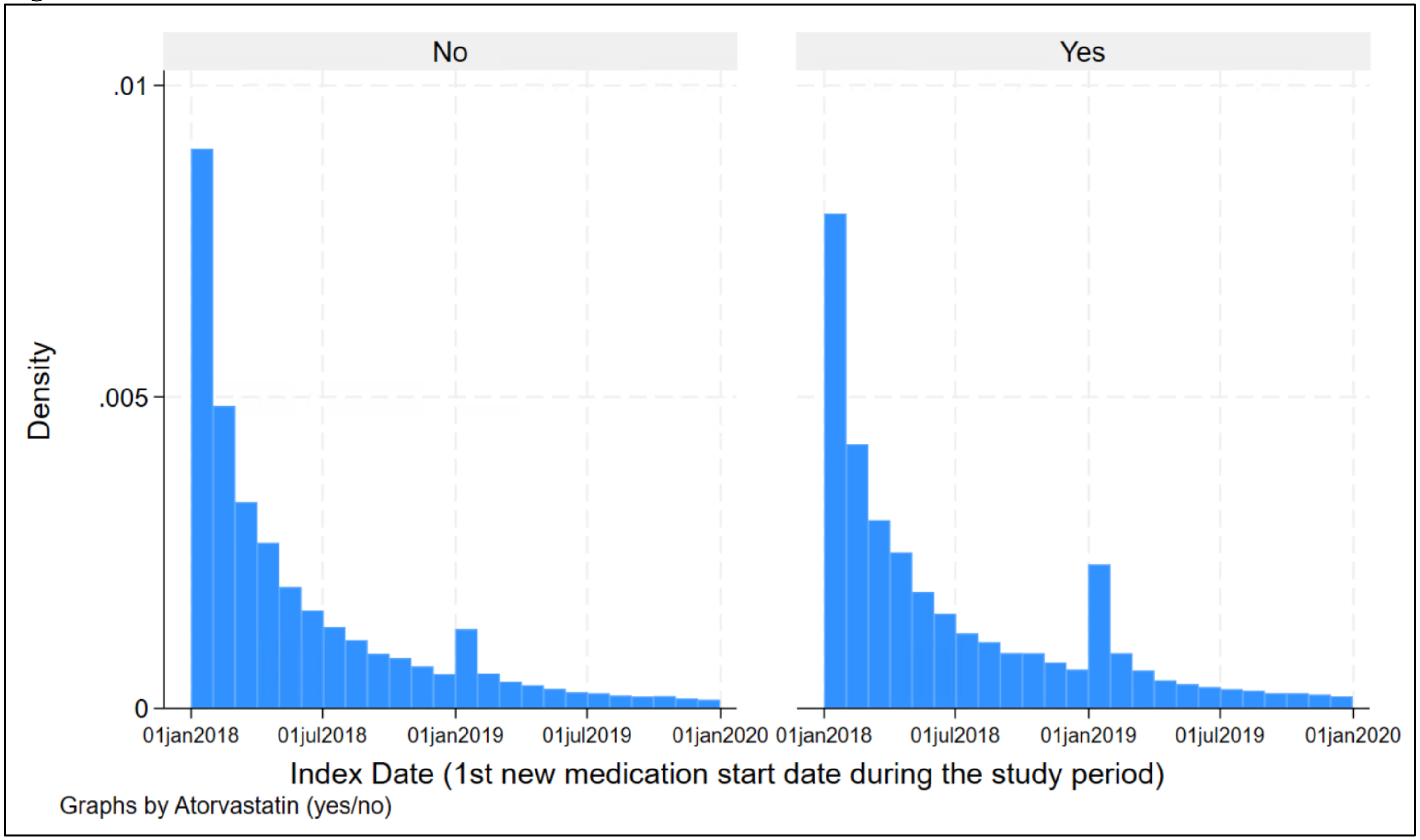

**Figure S2 Sensitivity Analysis - Distribution of Index Dates for Atorvastatin Initiators and Non- Atorvastatin Initiators**

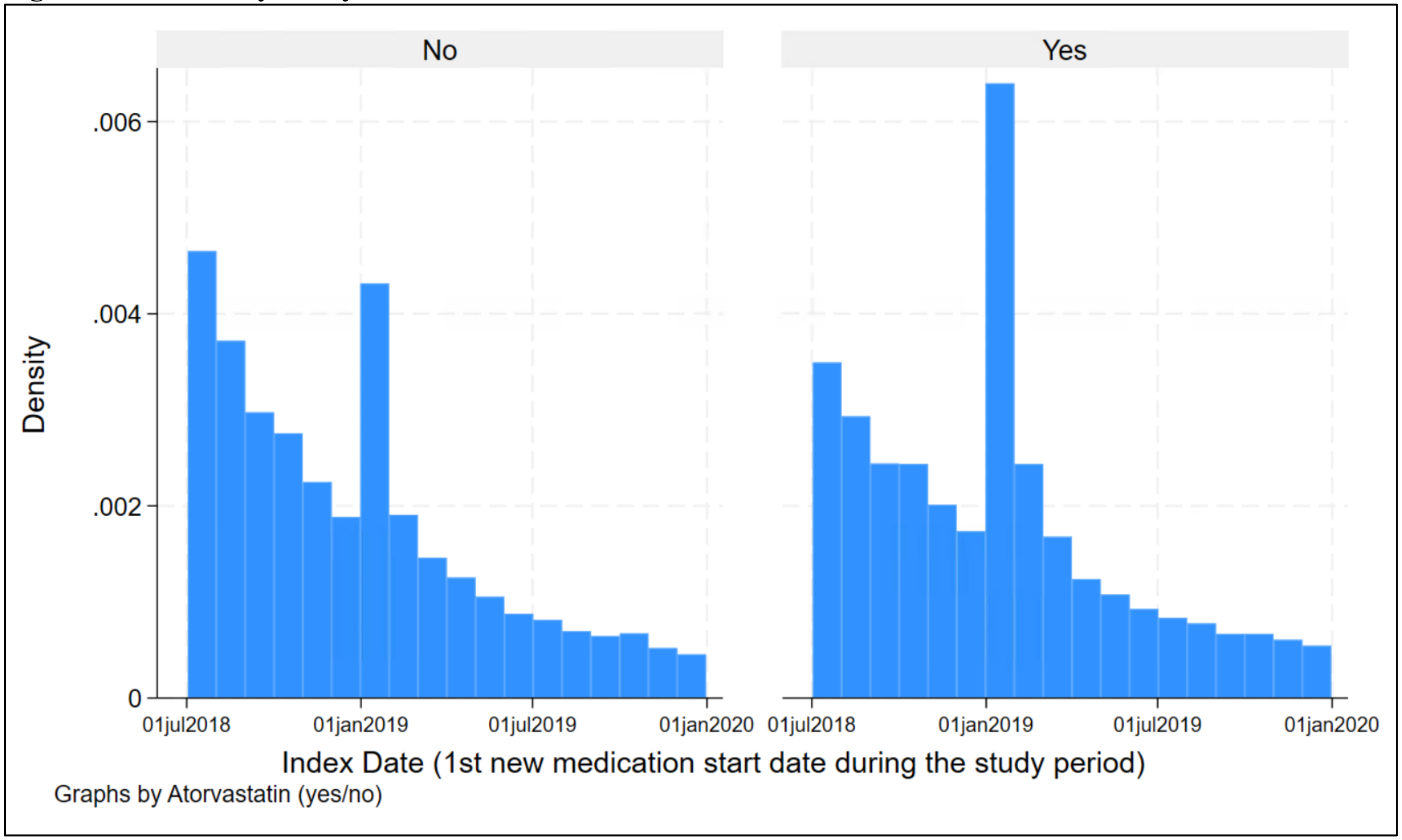
